# Explainable machine learning for post PKR surgery follow-up

**DOI:** 10.1101/2025.07.03.25330835

**Authors:** Corentin Soubeiran, Maëlle Vilbert, Benjamin Memmi, Cristina Georgeon, Vincent Borderie, Anatole Chessel, Karsten Plamann

## Abstract

Photorefractive Keratectomy (PRK) is a widely used laser-assisted refractive surgical technique. In some cases, it leads to temporary subepithelial inflammation or fibrosis linked to visual haze. There are to our knowledge no physics based and quantitative tools to monitor these symptoms. We here present a comprehensive machine learning-based algorithm for the detection of fibrosis based on spectral domain optical coherence tomography images recorded in vivo on standard clinical devices. Because of the rarity of these phenomena, we trained the model on corneas presenting Fuchs’ dystrophy causing similar, but permanent, fibrosis symptoms, and applied it to images from patients who have undergone PRK surgery. Our study shows that the model output (probability of Fuchs’ dystrophy classification) provides a quantified and explainable indicator of corneal healing for post-operative follow-up.

## 1. Introduction

During previous work, we have developed a human cornea analysis algorithm from Spectral Domain Optical Coherence Tomography (SD-OCT) images, based on the attenuation of the signal in the cornea volume[1,2]. This allowed us to distinguish healthy corneas from pathological corneas presenting Fuchs endothelial corneal dystrophy (FECD). However, this analysis is performed within a restricted, homogeneous area in the volume of the corneal stroma. Information on subepithelial fibrosis, symptomatic of advanced stages of FECD, was not exploited. This symptom of subepithelial hyperreflectivity in FECD is shared with another pathology: corneal haze, an inflammatory reaction, sometimes observed following Photorefractive Keratectomy, a form of refractive surgery. Clinicians do not have an objective, quantitative, physics-based and reliable tool for its evaluation and consequently encounter difficulties in diagnosing haze, especially in transient cases.

We present here an approach for the automatic classification of healthy and pathological corneas based on the morphological characterisation of the subepithelial fibrosis zone, visible in clinical OCT. After defining quantifiers of fibrosis, we use a machine learning model for the classification of clinical images showing Fuchs’ dystrophy and then its extension to the diagnosis of corneal haze. We show how this approach provides practitioners with a measurable and interpretable diagnostic tool.

## 2. Clinical assessment of corneal haze

Current means of assessing corneal haze are subjective rating scales (Fig. 1a)[3,4] and the measurement of corneal densitometry achieved with the Scheimpflug rotary camera, the average grey level of photographs provides an objectively characterising corneal scattering but without extracting physical parameters related to tissue properties (Fig. 1b)[5,6].

**Fig. 1.**
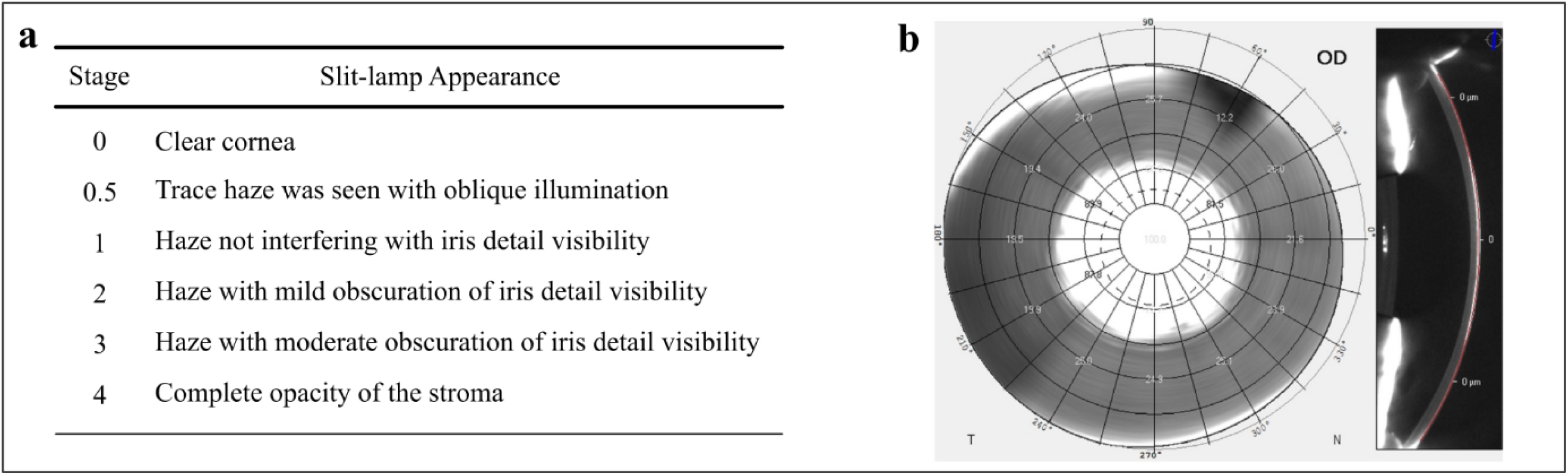
Clinical means for corneal haze evaluation. **(a)** Arbitrary scale for haze degree evaluation with slit-lamp reproduced from Ref. 3. **(b)** Densitometry map based on Pentacam© Scheimpflug images used as corneal scattering characterization, reproduced from Ref. 6.

## 3. Materials and methods

We chose a simple automatic classification approach using machine learning instead of the latest deep-learning approaches using convolutional neural networks. This approach aims to use interpretable parameters for the classification and therefore provide insight into the reason for the classification to clinical practitioners, for whom the consideration of such parameters in their clinical practice is facilitated if they do not perceive a “black box” aspect in the processing of the data.

### 3.1. Morphological parameters of subepithelial fibrosis

Compared to a healthy cornea (Fig. 2a), the subepithelial fibrosis of corneal haze and some FECD cases is characterised by increased light scattering due to mature myofibroblasts and gaps between type III collagen fibrils. Its presence gives rise to a perceived “hyperreflective” area in the anterior stroma (Fig. 2b and c).

**Fig. 2.**
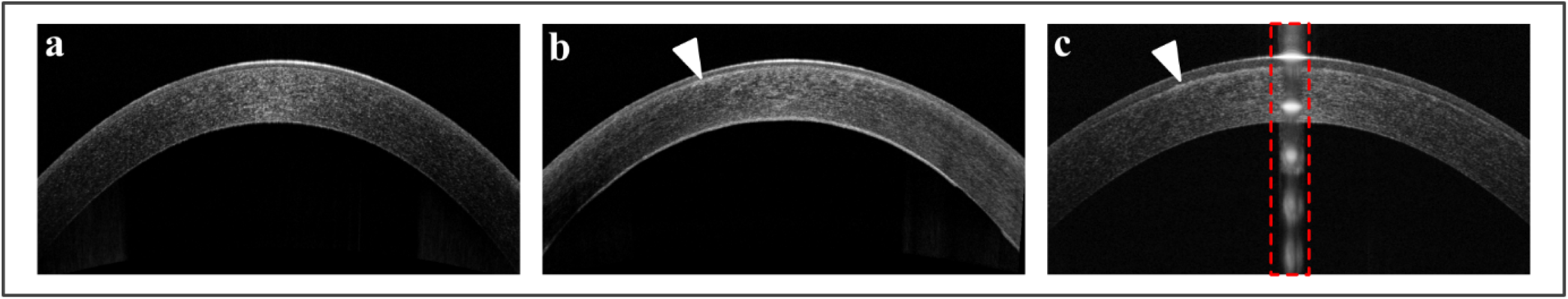
Comparison of SD-OCT images of **(a)**: a normal cornea, **(b)**: a cornea presenting Fuchs endothelial corneal dystrophy; the zone of subepithelial fibrosis is indicated by the arrow, **(c)**: corneal haze, the red doted square delimitates specular reflection an artefact of SD-OCT signal not to consider, the arrow indicates subepithelial fibrosis. Images have been recorded using the clinical SD-OCT RTVue-XR Avanti.

Ophthalmologists point out that this area of fibrosis distorts the automatic corneal layer segmentation algorithms built into clinical devices, providing an indirect clue to the presence of corneal haze: the hyperreflective area causes an overestimation of the epithelial layer thickness in the automatic detection, which the corneal thickness mapping algorithms interpret as epithelial hyperplasia[7].

We used the existence of these marked morphological features, already described qualitatively in terms of reflectivity profile in confocal images[7], to extract objective parameters for characterising subepithelial fibrosis from OCT images. The aim is to use these quantifiers as input parameters to establish a model for the automatic classification of clinical images. One of our major concerns has been to ensure the interpretability of these parameters by physicians.

### 3.2. Standardisation of clinical images

The SD-OCT images are flattened and standardised following the processing chain described in Ref. 2. This pipeline is supplemented with an additional alignment procedure relying on normalised cross-correlation. Each A-scan (OCT axial signal) is aligned according to the first air/lacrymal interface. If necessary, the central saturation artefact (specular reflection illustrated in Fig. 2c) is detected and removed from the image, the corneal image is digitally flattened and laterally normalised, and the posterior stromal artefact is compensated if detected by Principal Component Analysis (PCA). Low signal intensity regions on each side of the image are removed. Each parameter is estimated on an averaged depth profile of window size 1.5mm for each possible position of this window. The final value of the estimated quantifier is the weighted average for each window, the weight being the normalised distance to the centre of the cornea (minimal weight 0.5, maximum: 1). This procedure aims to reduce the impact from artefacts of the flattening procedure, and limited quality of OCT signal on the sides due to corneal curvature.

### 3.3. Definition of subepithelial fibrosis quantifiers

The morphological characterisation of subepithelial fibrosis is based on the identification of peaks of interest in the depth-averaged intensity profile of the corneal OCT signal: among others, the first peak corresponds to the air/tear film interface and the second to the epithelial basement membrane/Bowman’s layer interface (Fig. 3a-c). This second peak constitutes our signal of interest because the excimer laser acts from this depth onwards for the corneal stroma remodelling, at about 100 µm depth in the cornea. Therefore, at about this depth, the scarring reaction may lead to inflammation or even fibrosis, as shown in OCT by the deepening of the peak in question (Fig. 2c).

**Fig. 3.**
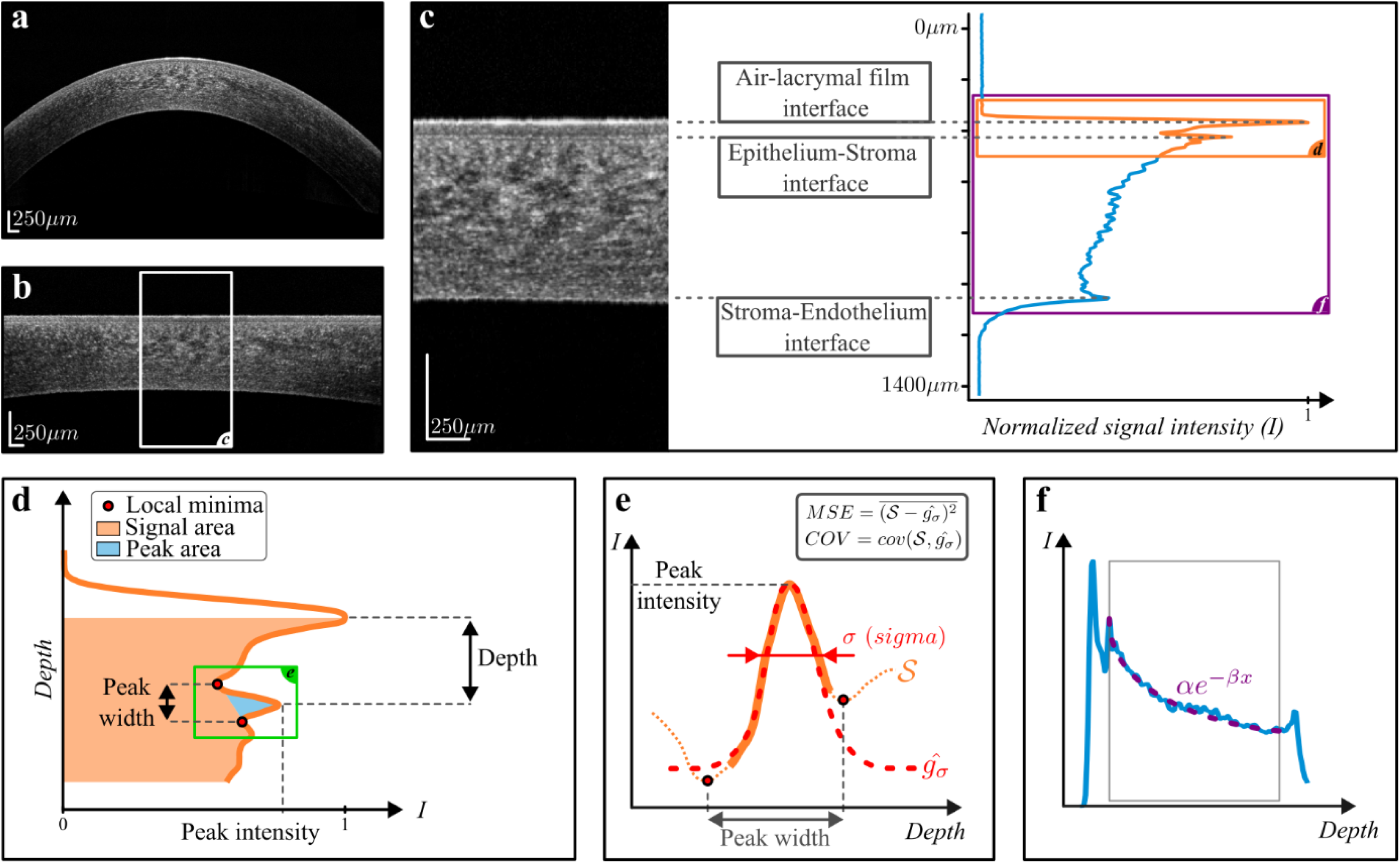
Extraction of morphological quantifiers associated with the epithelium/stroma interface performed on a healthy cornea OCT signal. **(a)**: OCT of a normal cornea acquired with clinical SD-OCT RTVue-XR. **(b)**: The standardized OCT image, limited to the central part of 6 mm (for an overall field-of-view of 8 mm during acquisition). **(c)**: 1.5mm window and associated laterally averaged depth intensity profile. **(d)** Zoom on the two anterior intensity peaks and definition of several morphological quantifiers for the peak associated with the interface between Bowman’s layer and the corneal stroma: Depth, Peak intensity, Peak Width, and peak Area Ratio. **(e)** Additional quantifiers to this peak resulting from profile estimation: Sigma, MSE and COV. **(f)** The last two quantifiers: Alpha and Beta corresponding to stromal signal decay.

The intensity peak associated with the epithelial basement membrane is identified via the two local minima around the peak (Fig. 3c,d). It is defined by its location in depth and delimited by the distance between the axial position of the peak and the location of the two local minima (Fig. 3d).

An interpolation is performed to improve the sampling in this area, and a Gaussian regression is applied to the intensity profile corresponding to the peak. The Gaussian regression provides four quantifiers: (Fig. 3d,e)

1. The **“Depth”** below air-lacrymal film interface(µm) to the estimated mean of gaussian regression.
2. Standard deviation “**Sigma**” of the peak: quantifies the spread of the peak in depth (µm). In the case of fibrosis, it therefore characterises its depth extent.
3. Distance between the two local minima “**Peak width**”: it also characterises the spread of the peak in depth but by looking at local minima independently to peak shape.
4. The maximum intensity of the second peak **“Peak Intensity”** compared to the maximum intensity of the first peak. It therefore characterises the relative intensity of the backscatter at the epithelium/stroma interface compared to the reflection at the air / lacrymal film interface.

Two parameters quantify the quality of the Gaussian regression: (Fig. 3e)

5. The mean square error **“MSE”**: characterises the deviation between the regression and the actual values of the profile.
6. The covariance **“COV”** between the profile and the regression: this quantifies the joint difference between the expectations of the profile and the Gaussian.

These last two parameters were defined following the observation of the images, and in particular, the observation that the Gaussian is sometimes poorly resolved for pathological corneas. High values of squared error or poor covariance can be a warning of pathology.

An exponential regression is then performed between the first peak and the endothelium to characterise the attenuation of the OCT intensity as it propagates through the stroma (Fig. 3f). Two additional quantifiers correspond to the fit variables of the regression function *f(x)* = *a* exp(―β*x*) + C with

7. the amplitude *a* **“Alpha”,**
8. the rate of decay β **“Beta”**.

Finally, a last quantifier characterises the optical contrast by the ratio of the area under the intensity profile curve with and without the second peak, removed by linear fitting between the two local minima on either side of the peak:

9. **“Area ratio”** (Fig. 3d).

The computation of these 9 quantifiers is coded in Python with a class architecture, which allows fast processing of a large number of images as objects and their attributes. The processing results for all images are accessible simultaneously.

The separation of quantifier distributions for a group of healthy corneas (166 OCT images) and a group of corneas with FECD (313 OCT images; inclusion criterion is selection for endothelial grafting, no stage of pathology specified) is statistically significant for 7 of these quantifiers: Peak Width, Sigma, Depth, MSE, Area Ratio, Alpha and Peak Intensity. (Fig S1)

### 3.4. Databases

Our final goal is an automatic classification of clinical OCT images into two categories: “healthy corneas” and “corneal haze”. Due to the difficulty of clinical diagnosis of haze, especially in mild cases, there are not enough validated clinical images to constitute a sufficient training database.

Therefore, we adopted the following strategy: train a classification model on a condition with similar symptoms and apply the same model to images with corneal haze. The pathology we focused on was FECD. In its advanced stage, it is characterised by fibrosis localised in the anterior stroma; the 9 morphological quantifiers previously defined are therefore a priori suitable to describe this pathology. Contrary to visual haze, FECD led to permanent symptoms, leading to an extended data collection.

The databases we exported (with patient or their legal guardian’s informed consent, see *consent and ethical considerations* section) from the RTVue-XR Avanti clinical SD-OCT devices.to feed this classification approach are listed in Table 1.

**Table 1.**
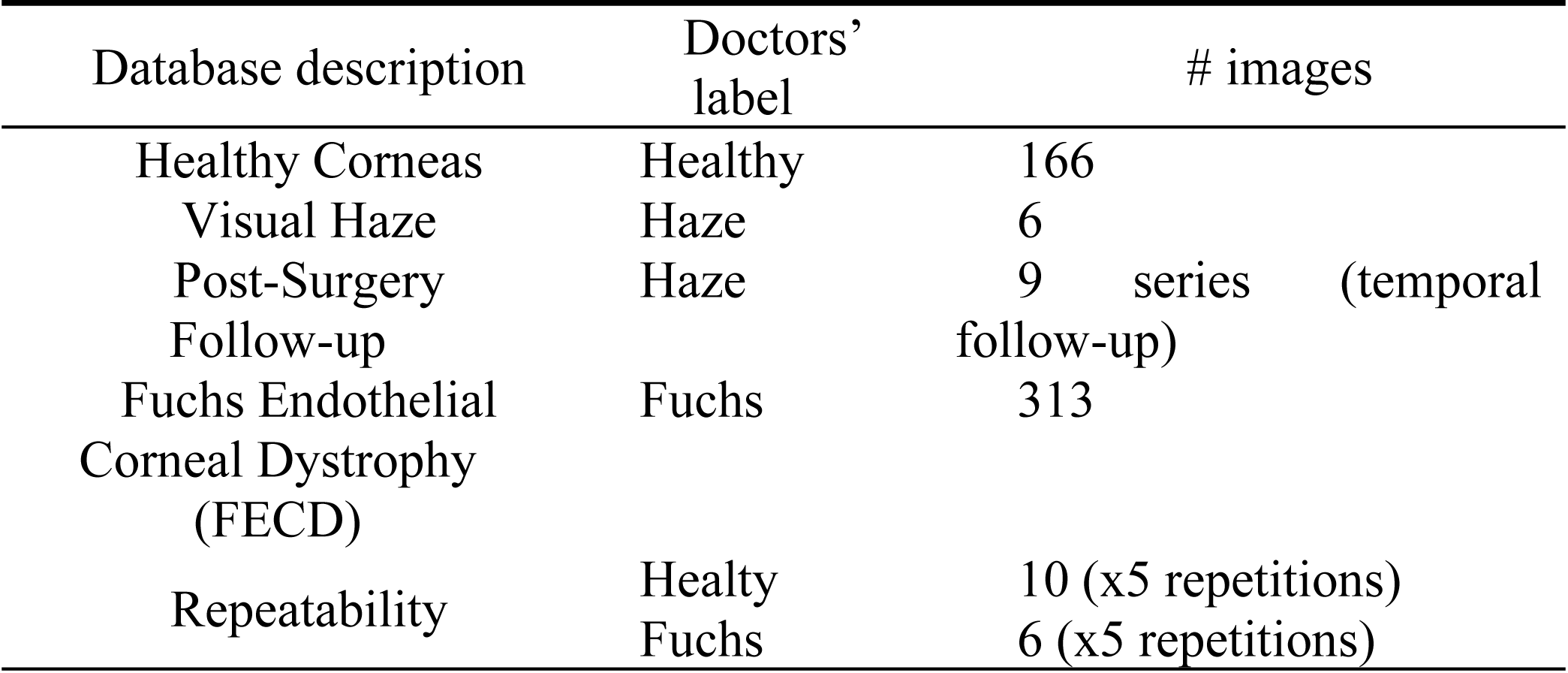
Databases used for the automatic classification exported from the RTVue-XR Avanti clinical SD-OCT. The pathologies of interest (corneal haze and Fuchs dystrophy) have in common the development of subepithelial fibrosis in their advanced stages.

### 3.5. Learning from corneas with Fuchs’ dystrophy (FD)

As a method to evaluate haze, we trained a model we trained on corneas presenting FECD. A first “one-class SVM” (Support Vector Machine) approach[8], which consists of describing a class (here the healthy cornea class) very precisely to confront it with healthy or pathological images, proved unsatisfactory results: the pathological corneas that were confronted with this characterisation were comparatively classified as healthy with more than 99% chance. We therefore randomly separated the databases of 166 healthy corneas and 313 corneas with FECD into two sets to train classical classification models: a training set, comprising 80% of the data, and a testing set with the remaining 20%, both maintaining class distribution for robustness[9,10].

The input parameters of the classification models are the morphological quantifiers; the output parameter is the **probability of FECD classification P_FECD_** predicted by the model, which indicates the degree of confidence given to the classification. A cornea is considered healthy if *p* < 50% and pathological if *p* > 50%.

The choice of classification model was evaluated by 5-fold cross-validation. Iteratively training different models on 4/5ths of the data and then validating on the last 1/5th. Cross-validation gives an idea of the accuracy of the models in the general case, without favouring one of the over-learned models, and allows the model selection for testing. Among the classification models tested (SVM-Linear, SVM-RBF, logistic regression, decision tree, random-forest, neural network), Random Forest proved the best result in validation and has been selected for testing (Table S1). Test accuracy was 96.9% with a classification of both specific (96.8% true positive rate) and sensitive (97.0% true negative rate) resulting in a ROC AUC of 0.9976 (Fig. 4a-c). The ambiguous cases of the three misclassified images will be discussed. The distribution of classification probabilities by the model on the testing set for the normal (33/166 images), Fuchs’ dystrophy (63/313 images) and Haze (6 images) databases is shown in Fig. 4a, and shows a significant separation between the control (normal corneas) and pathological (Fuchs dystrophy and haze) groups. This result justifies the rarity of ambiguities and underlines the interest in studying the few misclassified images.

**Fig. 4.**
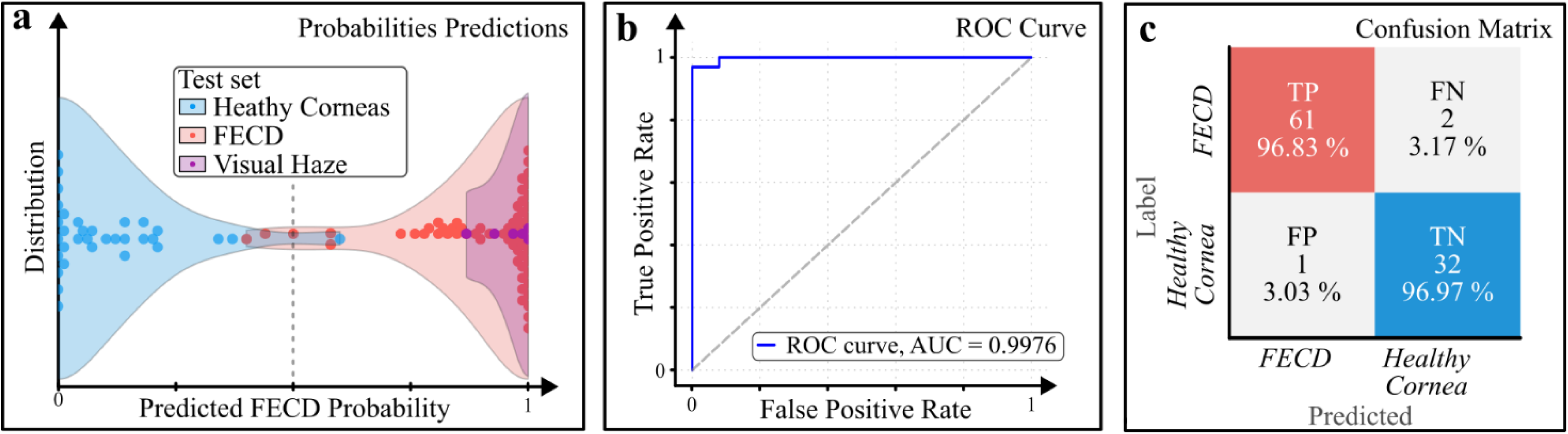
Random Forest Results on test dataset. **(a)** distribution of predicted dystrophy probabilities (Healthy mean predicted probability 10.0% std: 13.2%; Fuchs Dystrophy mean predicted probability: 90.9% std: 14/2%; Haze mean predicted probability 96.0% std: 4.7%). **(b)** Receiver Operating Characteristic (ROC) Curve, Area Under the Curve (AUC) 0.9976. (c) Confusion matrix.

### 3.6. Interpretability

As our method is grounded in physical parameters derived from the corneal OCT intensity signal, feature analysis provides direct insight into the decision-making process of our machine-learning model. To achieve this, we employed SHapley Additive exPlanations (SHAP)[11], a widely used framework for interpreting machine learning predictions. SHAP is model agnostic, meaning it can be applied to any machine learning model regardless of its internal structure, including ensemble methods, neural networks, and linear models. SHAP quantifies the contribution of each input feature to a model’s output by computing Shapley values, a concept from cooperative game theory. These values represent the marginal contribution of a feature to the prediction when considered in all possible subsets of features. For a given feature *i*, its Shapley value ϕ_i_ is computed as (adapted from Ref. 11):

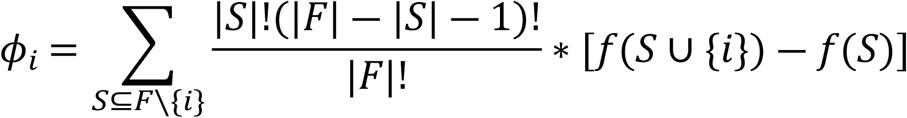

where *F* is the set of all features, *S* is a subset of features excluding *i*, and *F(S)* is the model’s output based on *S*. In practice, SHAP approximates *f(S)* and *f(S ∪{i})* without retraining the model for every subset. Instead, it marginalizes the model’s predictions over the distribution of missing features. For tree-based models, the TreeExplainer variant computes these values efficiently by leveraging the tree structure to conditionally traverse decision paths and split probabilities.

This approach ensures a fair allocation of the predicted outcome among all features, satisfying properties of consistency and additivity. SHAP provides both global interpretability by aggregating feature contributions across the dataset and local interpretability by explaining individual predictions. These capabilities allowed us to identify the most influential features driving the model’s decisions while ensuring transparency and alignment with the underlying physical phenomena.

**Fig. 5.**
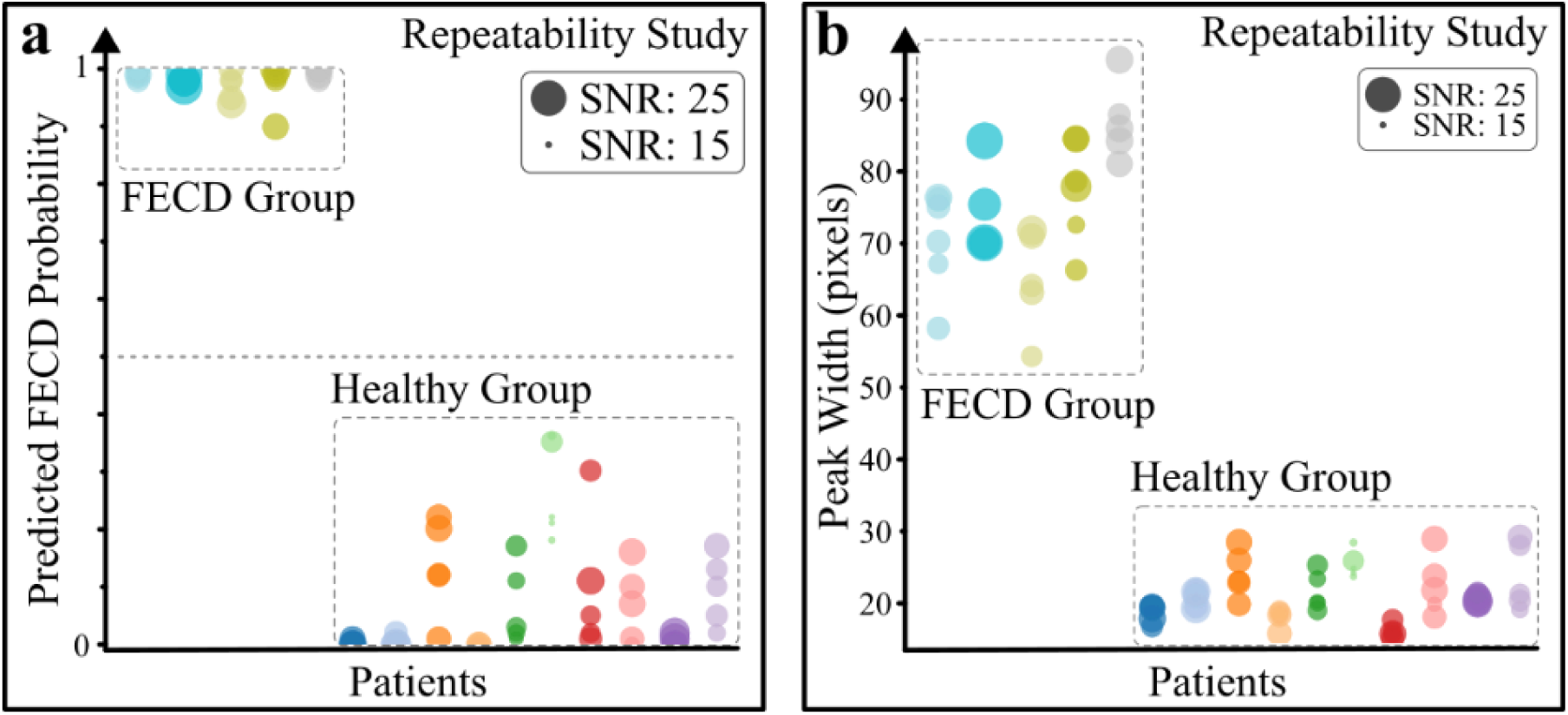
Results of the repeatability study on 10 normal corneas (group control) and 5 corneas with Fuchs’ dystrophy (pathological group). **(a)**: Probabilities of dystrophy classification for the 5 images per corneas, respectively normal and pathological. **(b)**: Repeatability of peak width quantifier.

## 4. Results

The robustness of the trained artificial intelligence model for the classification of healthy corneas and corneas with FD is demonstrated, with an accuracy of 96.9% and repeatable classification. The objective of our approach is to confront this same model with corneas that have undergone refractive surgery by PKR and developed a corneal haze, which is also manifested by subepithelial hyperreflexia.

### 4.1. Repeatability

A repeatability study was performed to test the robustness of the classification. We tested the model on a series of 5 images acquired on the same day by the same orthoptist on the same patients, including 10 healthy subjects (10 corneas) and 3 patients with Fuchs dystrophy (5 corneas). The results of the repeatability study are shown in Fig. 5. The classification accuracy of the model is 100%, with a compact and statistically significant separation between the healthy and pathological groups. However, we denote a strong dispersion of results in the Healthy group (Fig. 5a.) that do not correlate with the SNR index, nor with quantifiers dispersion as illustrated with peak width in Fig. 5b. Nevertheless, we noticed variation in the OCT signal itself in terms of signal intensity, texture or artefacts, as illustrated in Fig S2.

### 4.2. Validation

As a first step, we validated the extension of this model to a small database of 6 corneas identified with haze by specialists. The 6 corneas have been classified in the dystrophy group without additional training, leading to a 100% detection rate (average predicted FD class probability: 0.960, std: 0.047).

### 4.3. Post-surgery follow-up

To evaluate the potential of our approach for the postoperative follow-up of refractive surgery by PRK, we collected OCT images of the clinical follow-up up to 8 months of 5 patients operated by refractive surgery (9 operated corneas). The inclusion criterion was the diagnosis of a post-operative corneal haze, without any criteria for the severity of inflammation: the scarring reaction may therefore be mild.

The quantitative follow-ups, shown in Fig. 6a, demonstrated for all patients that at least one occurrence was classified in the FECD class after surgery (FECD probability >0.5). Some patients are classified in the FECD class before surgery too, PRK being sometimes performed on pathological corneas. We found it insightful to represent this follow-up relatively to the initial value of the probability and therefore represent potential improvement or degradation of such quantifier (Fig. 6b). One can then distinguish corneas classified as not FECD (FECD probability <0.5) before the surgery have lowered probabilities after surgery at least for a temporary period. And corneas initially classified as pathological are trending toward an improvement in the quantifier.

**Fig. 6.**
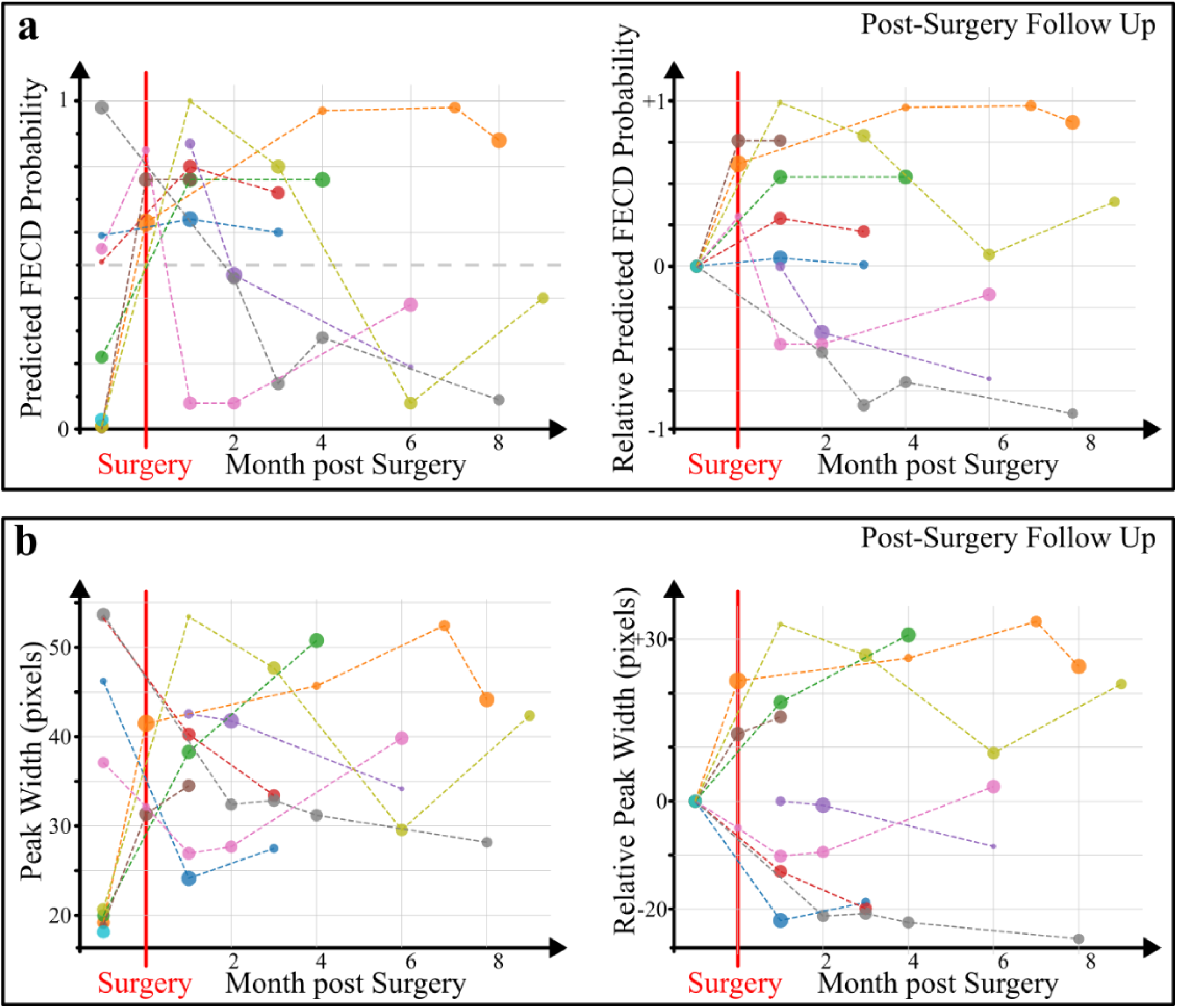
Haze post-surgery follow-up study **(a):** Probabilities (left) and relative to initial probability (right) of dystrophy classification in Pre/Post-surgery follow-up for 9 patients. **(b)** Pre/post-surgery follow-up of peak width. The size of the points is relative to the signal-to-noise ratio of the OCT image analyzed

### 4.4. Interpretability

By aggregating the absolute Shapley values across all instances, we derived a ranked feature importance profile (Fig. 7a), highlighting the peak width as the most influential variable contributing to the model’s predictive accuracy. The SHAP summary plot (Fig. 7a.) revealed nuanced insights into feature interactions: for instance, higher values of peak width, sigma, area ratio, and depth; and lower values of beta (reduced stromal intensity signal decay) are associated with higher SHAP values and, therefore, a higher FD class probability prediction. The feature importance deduced by SHAP is confirmed by traditional impurity-based importance scores (e.g., Gini Impurity) (a higher score indicates that the feature contributes more to the predictive power of the model) (Fig. 7b.). Additionally, SHAP provides a local explanation, for each prediction it shows how each feature either increases (pushes toward) or decreases (pushes against) the predicted value compared to a baseline (e.g., the mean prediction or expected value). For instance, on a true healthy cornea, all features participate in the predicted probability of 0 (Fig. 7c, top); conversely, on a true FECD cornea, all features participate in the predicted probability of 1 (Fig. 7c, bottom). In those two examples, Peak width, Sigma, and area ratio are the main participating features. Additionally, such a study can be made on misclassified data to understand what leads to a wrong prediction (Fig. 7d, e) and will be discussed later. Altogether, these pieces of information indicate the peak width as a potential good biomarker of sub-epithelium fibrosis. Training the Random Forest model only with the peak width parameter leads to a classification accuracy on the FECD/healthy dataset (evaluated by cross-validation) of 88.5% (std: 2.94%), similar accuracy is found for SVM or logistic models. The use of the additional 8 quantifiers allows us to reach a testing accuracy of 96.9%.

**Fig. 7.**
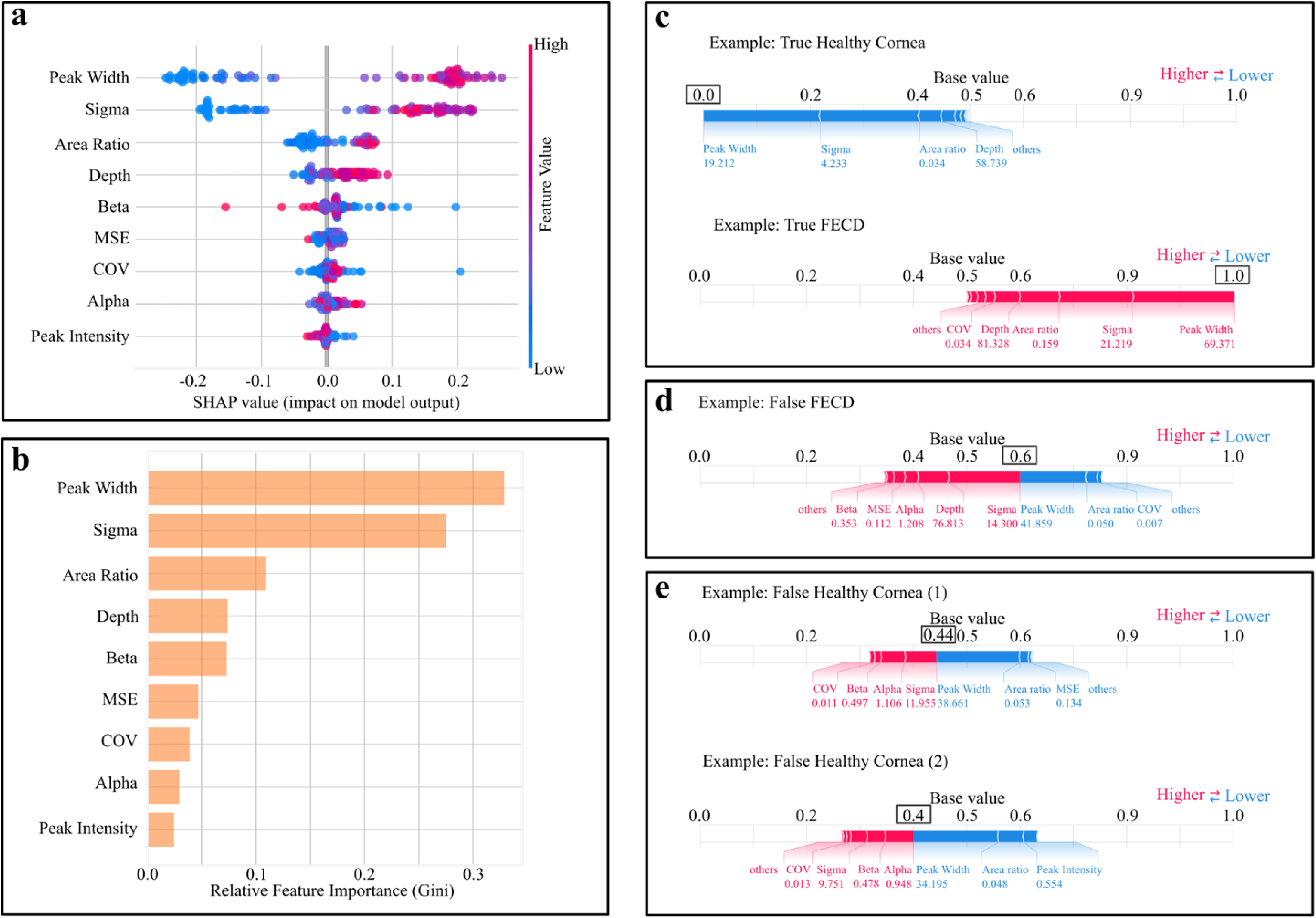
Interpretability insights. **(a)**: Shapeley Additive explanation (SHAP) features importance displaying how each training les (dots) influences the model output through lower SHAP values (tendency to healthy class) or higher SHAP values (tendency to Dystrophy class). Features are ordered from higher to lower impact (top to bottom). **(b)**: Gini feature importance. **(c)** Example of SHAP analysis for True Healthy Cornea (top) and True Cornea with Fuchs’ Dystrophy. The contribution of each feature to output prediction is illustrated. **(d)**: False Healthy cornea SHAP analysis. **(e)**: False Fuchs’ Dystrophy SHAP analysis.

This prediction model, combined with a SHAP analysis, could trigger clinicians’ attention to the physical aspects of the OCT corneal signal and help with post-operative follow-up as a complement to their expertise.

## 5. Discussion

### 5.1. Misclassifications

Although all haze images have been classified in the FECD class without re-training, one Healthy cornea and two corneas with FECD have been misclassified. First, it is worth noticing that the model for those 3 images outputs a probability close to 0.5 (Fig. 4a; Fig. 7d,e) denoting from the rest of the dataset predictions. An extended SHAP study on those three images reveals for the “False Fuchs Dystrophy” cornea a potential conflict between peak width and sigma quantifiers (Fig. 7d). Similarly, for the “False Healthy” corneas a conflict between peak width and sigma and stromal signal parameters (alpha and beta) is revealed (Fig. 7e). Average signals do not demonstrate particular features (Fig S3). Introducing new quantitative parameters sensible to signal density, disparity, or textural information might be relevant to identifying interpretation biases that are sources of error in image classification. Alternatively, convolutional neural networks (CNNs)[12,13] although less interpretable might detect some local fine features not caught by our approach. Additionally, using technics as gradCAM[14] could highlight directly on SD-OCT images the regions participating the most in the classification.

### 5.2. Post-operative follow-up after refractive surgery

According to the clinical data, transient haze is typically expected within 2-6 weeks post-surgery, maturing between 1-3 months and then regressing within a few weeks to 12 months [6]. There is indeed an increase in FD class probability p between 1 and 3 months post-op and either a reduction (improvement) or stagnation in 8 over 9 cases and only one case without improvements (see Results section and Fig. 6a).

Fig. 8 shows the criterion obtained by our approach in the case of mild corneal haze: it is difficult to distinguish between normal images (pre-surgery Fig. 8b and at M9 Fig. 8f) and corneas with haze (at M1 Fig. 8c) justifying how insightful this single FD probability indicator can be in post-op follow-up. However, the sample of corneas studied (9 eyes) is too small to draw any conclusions about the validity of this approach for fine haze tracking over time. The study should be extended to a larger and more temporally resolved sample to validate the present approach. A densitometry study shows the persistence of stromal diffusion more than twenty years after surgery for procedures with a large ablation depth[15]: it would be relevant to compare the model estimate for similar cases.

**Fig. 8.**
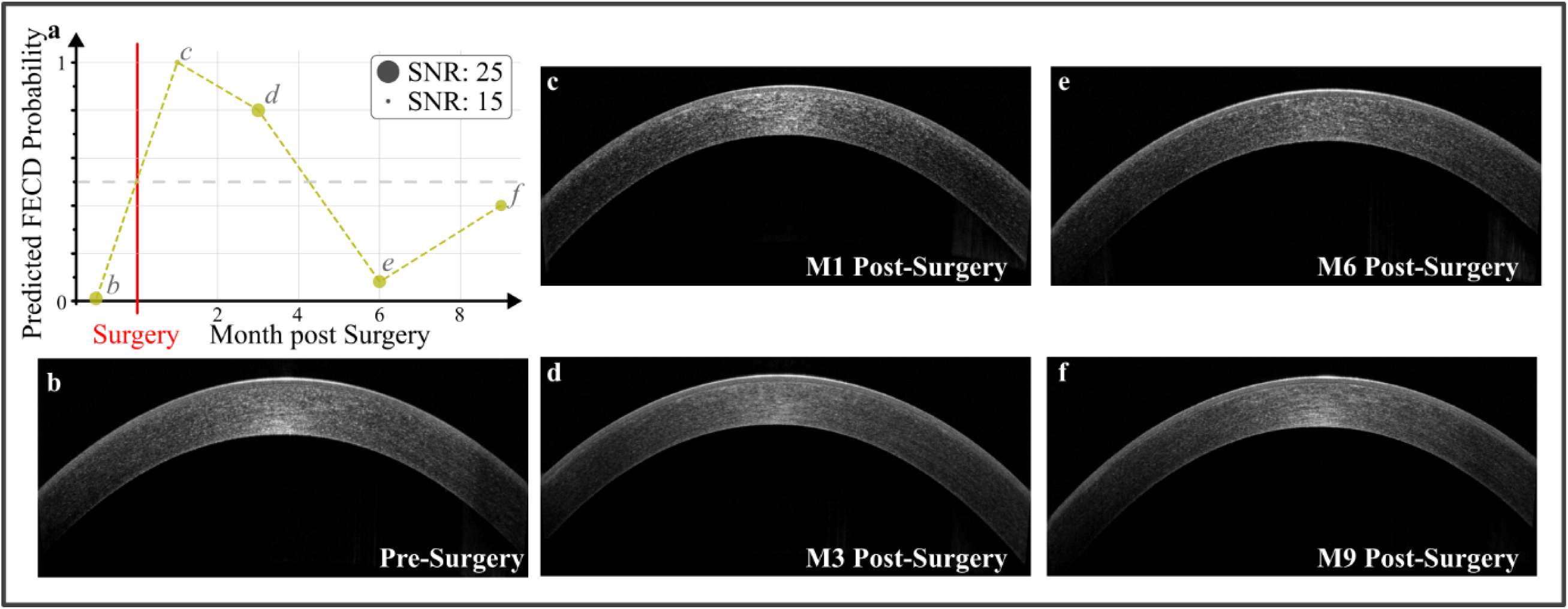
Example of surgery follow up. **(a)**: Evolution of Fuchs Dystrophy probability predicted by the model used as an indicator of fibrosis pre and post-surgery. **(b-f)** associated SD-OCT images recorded respectively before and 1,3,6,9 month after surgery.

### 5.3. Diagnostic tool

Obtaining a single criterion at the output of the model, in this case, the FECD probability classification p of the OCT images, is to be considered as one indicator among others in a diagnostic aid context. The probability p reflects the degree of confidence in the classification: in the case of a very clear-cut value, close to 0 or 1, the ophthalmologist has a good chance of being able to classify the image. Classification of images of healthy and pathological corneas clear confirmation of his clinical findings (extreme cases are generally well interpretable with a slit lamp); values close to 0.5 (or going against the expected classification) are a warning to ophthalmologists. In this second case, the other quantifiers are potential indicators of pathology, for example: alpha and beta factors giving information on signal decay or MSE, which reflects the Gaussian or non-Gaussian aspect of its depth profile, SHAP analysis helping to prioritise features to analyse on a specific example as described in Fig. 7c-e.

It would be tempting to rely on the peak width quantifier alone for clinical diagnosis, given the 88.5% classification accuracy it provides in validation. However, this is not sufficient: although this parameter has a strong impact on the model and has a high interpretability in terms of the axial extent of fibrosis, a classification accuracy of 88.5% is below the a-risk commonly set at 95% in medical statistics. Using the full set of 9 quantifiers allows us to exceed this threshold, confirming the clinical viability of our approach in terms of classification accuracy.

## 6. Conclusion

In this paper we propose a robust approach for automatic classification of clinical SD-OCT images for the detection of possible subepithelial fibrosis. The model, based on 9 morphological quantifiers and trained on Fuchs dystrophy, is 96.9% specific, sensitive and reliable. We demonstrated how the use of a common disease (in this case FD) helps to train and apply machine learning to a rarer, temporary pathology (haze) with similar symptoms. The transfer of the same model allows the detection of post-refractive surgery haze by PRK with 100% accuracy. A single criterion of “probability of FD classification” of an OCT image is provided by the model. It gives information on the degree of confidence associated with the classification. This criterion, which is explainable, is strongly related to the axial extent of fibrosis, which guarantees its interpretability by practitioners.

## Code and data availability

The codes associated with this approach are freely available in the following GitHub directory: https://github. com/Coohrentiin/Quantification_of_corneal_surgery_antecedent_2025. For consent reasons, data cannot be shared and are not available.

## Consent and ethical considerations

This retrospective study was conducted between January 1, 2019, and January 1, 2023. The study adhered to the tenets of the Declaration of Helsinki and was approved by the Ethics Committee of the French Society of Ophthalmology (IRB #00008855). Informed consent was obtained from all participants or their legal guardians. Data were anonymised before processing.

## Data Availability

The codes associated with this approach are freely available in the following GitHub directory: https://github.com/Coohrentiin/Quantification_of_corneal_surgery_antecedent_2025. Raw data cannot be shared and are not available.

## Acknowledgments

This study was partially funded through the “Prix de la Banque Francaise des Yeux 2022” obtained by Anatole Chessel.

**Corentin SOUBEIRAN** is a PhD student in BioPhotonics at Laval University, Quebec Canada. He received an engineering degree from ENSTA, (Paris campus), France, in 2023 in Artificial Intelligence. His work mainly focuses on the application of artificial intelligence in the domain of medical imaging and microscopy.

## Supplementary Materials

**Fig S1.**
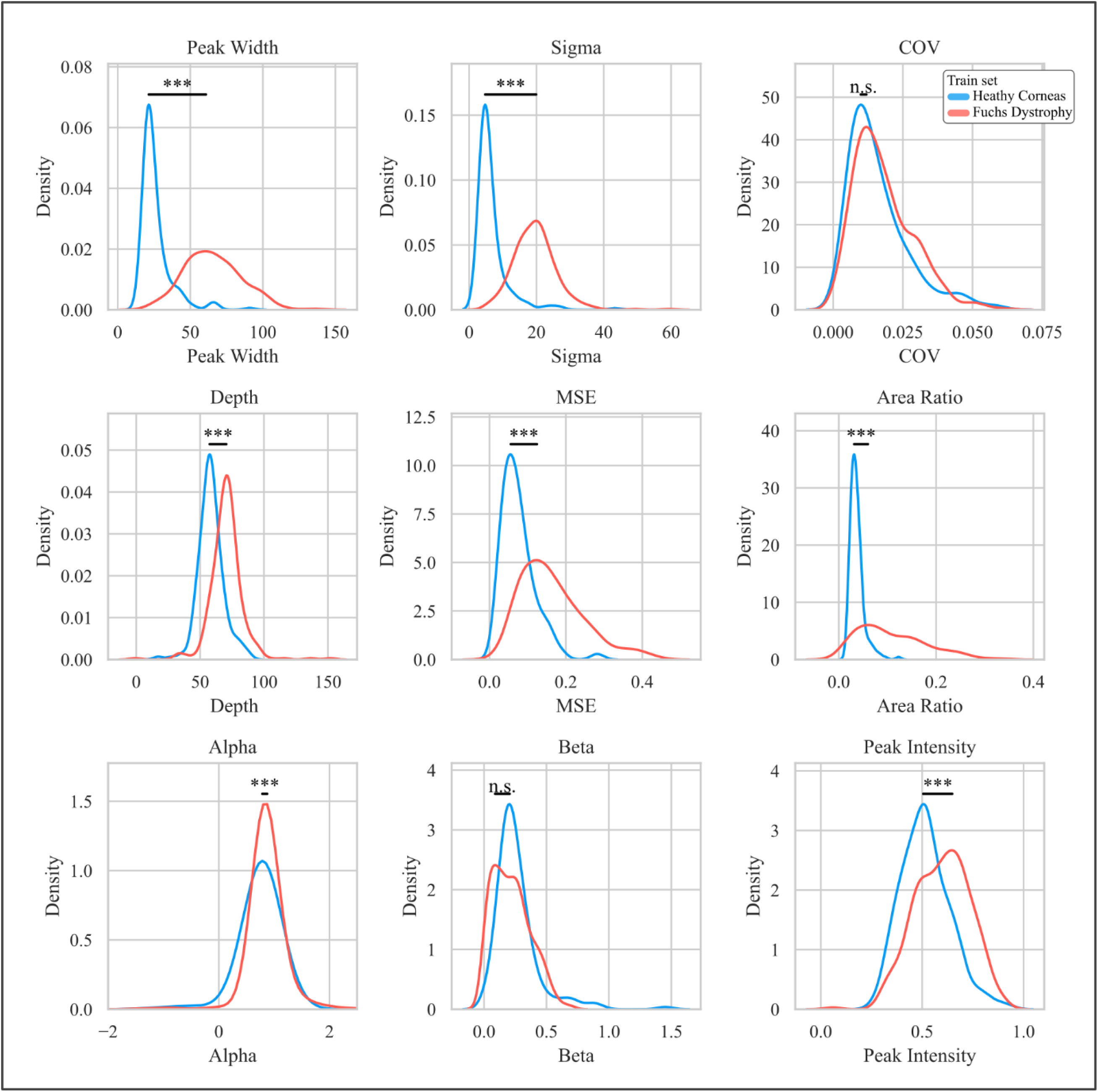
Distribution on training set of the 9 quantifiers for each condition. Statistics are performed with Mann-Whitney U-test (no normality assumption) using Bonferonni correction of p-values (***: p-value<0.001/m; **; p-value<0.01/m; p-value<0.05/m; m=9)

**Fig S2.**
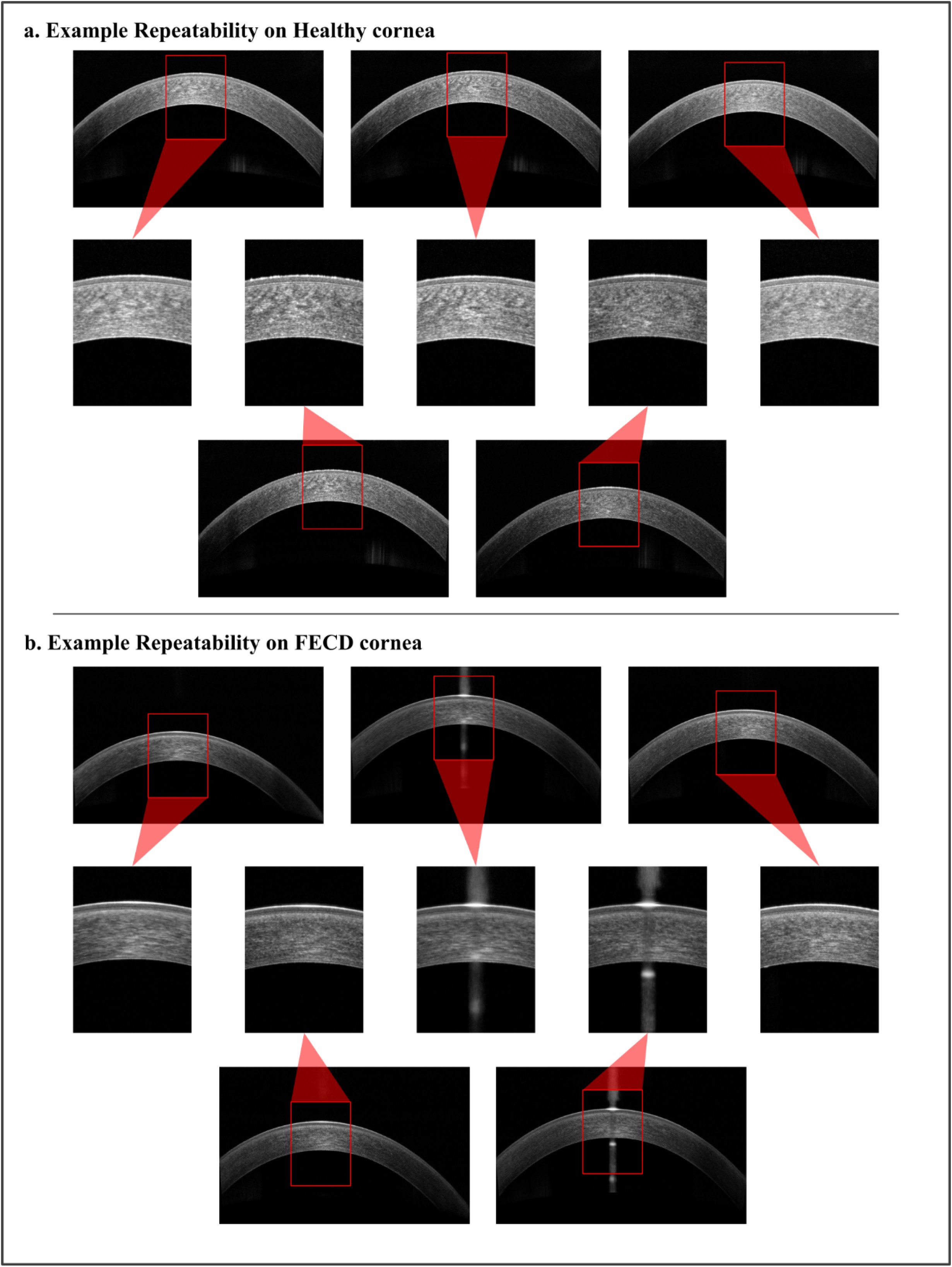
Raw Repeatability data example with zoom on the apex region **(a)**: healthy cornea example, **(b)**: FD cornea example.

**Fig S3.**
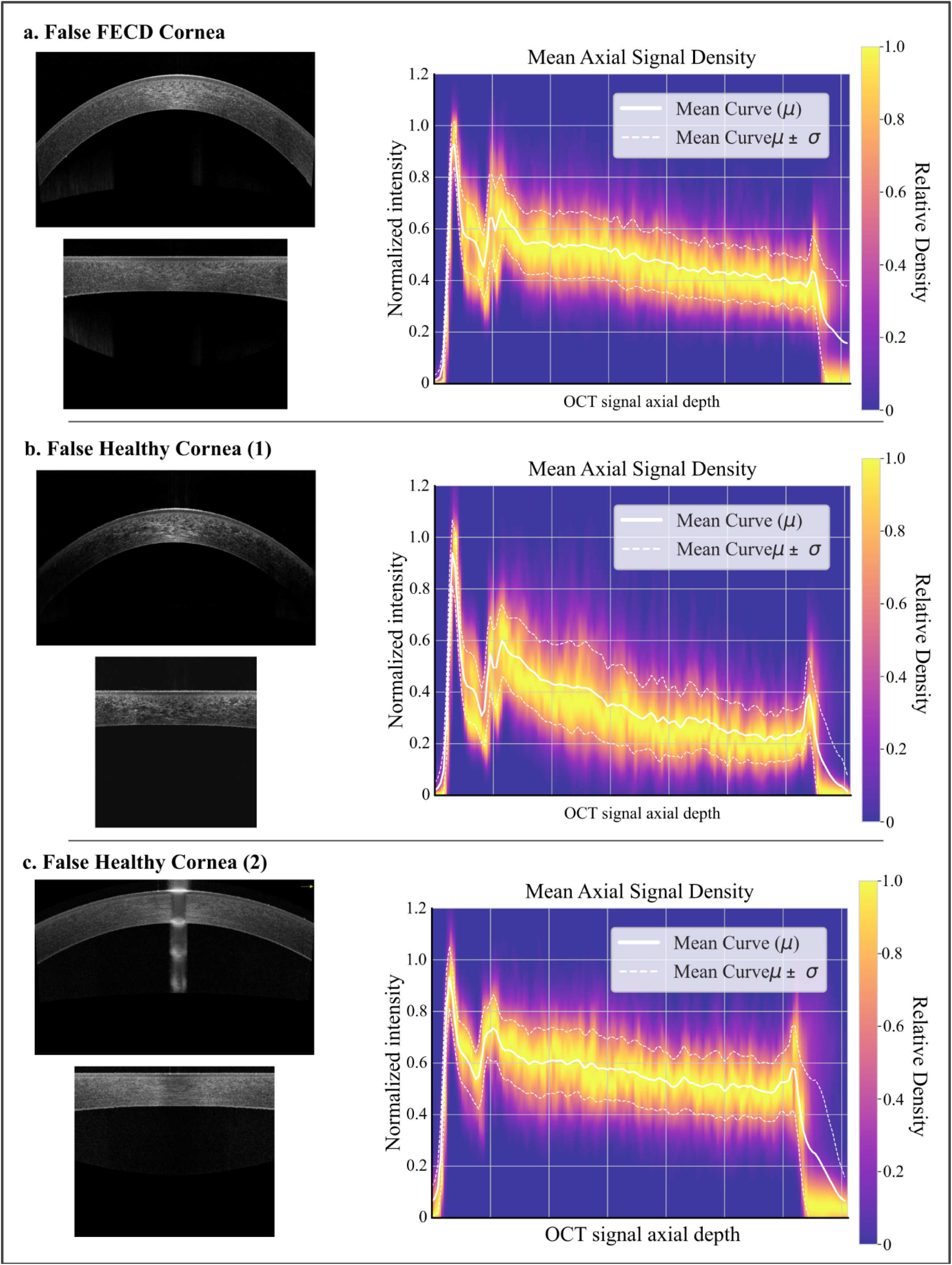
The 3 Misclassified corneas and corresponding SD-OCT raw signal (right top) associated standardized signal and depth profile (right) as a density map to represent signal disparities. **(a)**: false Fuchs dystrophy cornea. **(b-c)**: false Healthy cornea

---

**Table S1.**
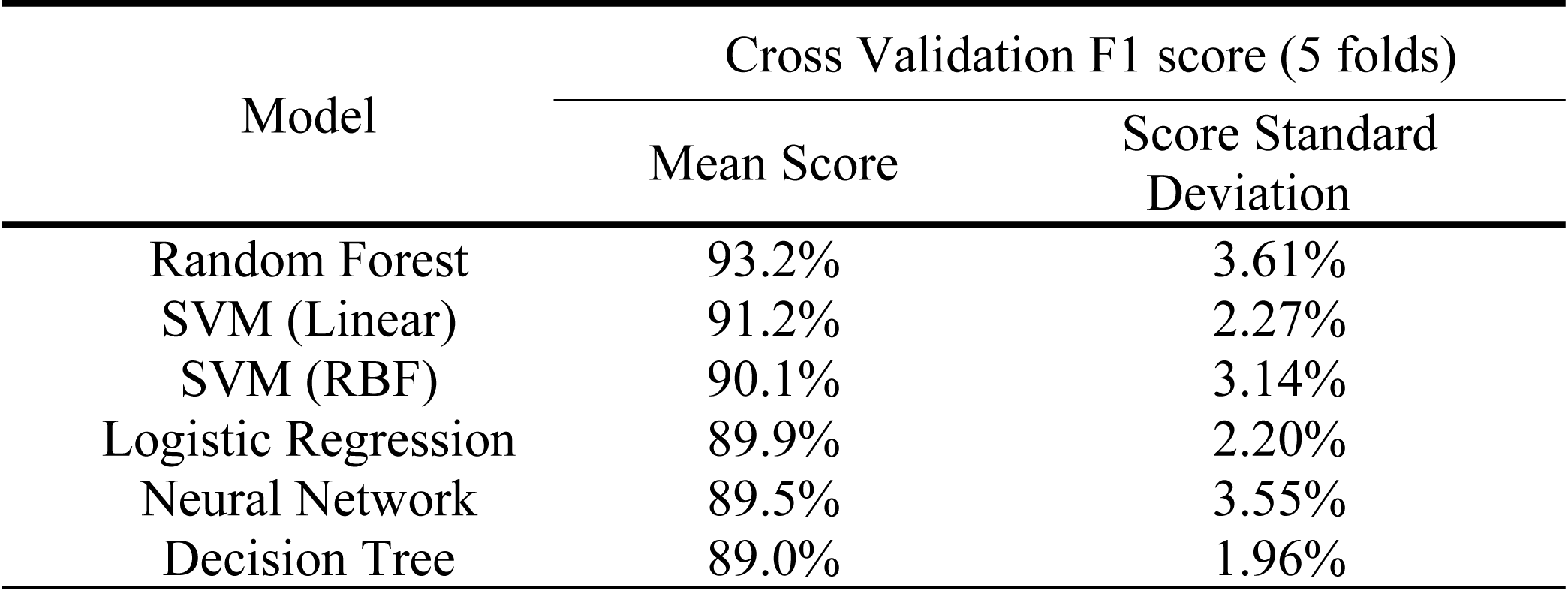
Cross-validation scores of classic machine learning classification models.

